# Overlooked bias with thermometer evaluations using quickly retaken temperatures in EHR: applications to axillary, oral, temporal artery, and tympanic thermometry

**DOI:** 10.1101/2020.11.24.20237958

**Authors:** Charles Harding, Marybeth Pompei, Dmitriy Burmistrov, Francesco Pompei

## Abstract

Studies related to COVID-19 increasingly use electronic health records (EHR) to obtain large-scale evidence. However, EHR-based research must be performed with care because it can involve new study design problems that are unfamiliar to much of the medical community. Haimovich et al. (2020) sought to inform COVID-19 practice by evaluating temporal artery thermometers (TATs). They retrospectively searched EHR for temperatures measured twice within 15 minutes, including once with a TAT. The TAT often disagreed with reference measurements, so Haimovich et al. concluded TATs perform poorly. Here, we extended Haimovich et al.’s study design to all other major thermometer types using the eICU Collaborative Research Database. We retrospectively identified 80,065 pairs of quickly retaken temperatures from 24,765 adult U.S. critical care patients treated in 2014-2015. We found that oral, tympanic, and axillary thermometers disagreed with reference measurements as much as TATs did. Moreover, all thermometer types showed unprecedentedly worse agreement than observed in research using other study designs: every thermometer type broke ±0.9°F (±0.5°C) limits of clinically acceptable agreement by >2-fold and no type satisfied basic standards for repeatability. A natural explanation for these findings is that clinicians often retook temperatures within minutes because of user or patient errors during measurement, such as probe misplacement or patient movement. This means that quickly retaken EHR measurements do not reflect device accuracy or precision in correct use and, contrary to Haimovich et al.’s conclusions, should not be used to evaluate thermometer performance or revise COVID-19 fever thresholds. Our study provides an illustrative example of unexpected study design problems that can undermine EHR-based research.

## Introduction

Studies related to COVID-19 increasingly use electronic health records (EHR) to obtain large-scale evidence. However, EHR-based research must be performed with care because it can involve new study design problems that are unfamiliar to much of the medical community.

Recently, Haimovich et al.^1^ sought to inform COVID-19 screening practices by evaluating temporal artery (forehead) thermometers (TATs). They retrospectively searched EHR for temperatures measured twice within 15 minutes, including once with a TAT. The TAT often disagreed with a reference measurement, so Haimovich et al. concluded TATs perform poorly.

Here, we extended Haimovich et al.’s analysis to other major thermometer types. We found that oral, tympanic, and axillary thermometers disagreed with reference measurements as much as TATs did. A natural explanation is that clinicians retook temperatures because they suspected a measurement had been taken incorrectly. This makes retaken EHR temperatures invalid for evaluating the accuracy and precision of correctly used thermometers, and also invalid for suggesting a revised COVID-19 fever threshold, which Haimovich et al. did. Our study provides an illustrative example of the unexpected study design problems that can undermine EHR-based research.

## Methods

All study data^2^ and code^3^ are available online. The eICU Collaborative Research Database v2.0 provides deidentified EHR from 2014-2015, including 139,367 intensive care unit patients at 204 US hospitals.^4^ In this retrospective, observational research, we studied 154 hospitals with ≥50 chart records listing studied thermometer sites. Excluding 4,852 patients aged <18 years and 1431 without temperature records, 108,970 adult patients and 5,304,829 temperatures were available. We excluded 30.4% of temperatures because a studied thermometer site was not listed, 0.3% as implausible (<50°F or >120°F), and 104 as potential double entries. From the remaining 3,670,376, we identified measurements retaken within 15 minutes: 160,130 matched temperatures (4.3%) from 24,765 patients. For measurements retaken repeatedly within 15 minutes, we included the first 2 temperatures only. Alternatively, one could average temperatures taken at the same site within 15 minutes,^1^ but that would make values less noisy for sites retaken more frequently, biasing Bland-Altman results.

Axillary, central, oral, temporal, and tympanic sites were studied. Each has strengths and weaknesses in terms of accuracy, safety, and convenience.^5^ Central temperature was defined as pulmonary artery, urinary bladder, esophageal, rectal, or core (subtype unspecified). Pulmonary artery is the only true gold standard,^6^ but is highly invasive and rarely taken (0.6% of temperatures), so all central temperatures were used as a reference standard instead. Site was recorded in 1-4 words of free text. We recoded entries appearing ≥10 times, amounting to 99.98% of entries.

We evaluated agreement between paired temperatures using Bland-Altman analyses. Analyses were cluster bootstrapped by patient to address within-patient nonindependence (replicates=20,000). Re-running analyses with only 1 measurement pair per patient also produced similar results.

## Results

Patients with quickly retaken temperatures had mostly similar characteristics to the overall patient population: 44.2% and 45.4% women, mean ages 65 and 64 years, 21.6% and 20.2% with diabetes, and 14.1% and 11.7% with admission diagnosis of sepsis. Paired temperatures (*n=*160,130) were 12.6% axillary, 45.4% central, 29.2% oral, 10.2% temporal, and 2.6% tympanic.

Every thermometer site had similar, very low agreement with reference temperatures (Figure 1). No site satisfied the ±0.9°F (±0.5°C) limits of agreement often used to define clinical acceptability.^6^ When analyzing temperatures retaken at the same site, every site showed unprecedentedly low repeatability (Figure 2). Retaken central temperatures were often anomalously cold (9.9% <95°F and 4.2% <92°F).

**Figure 1.**
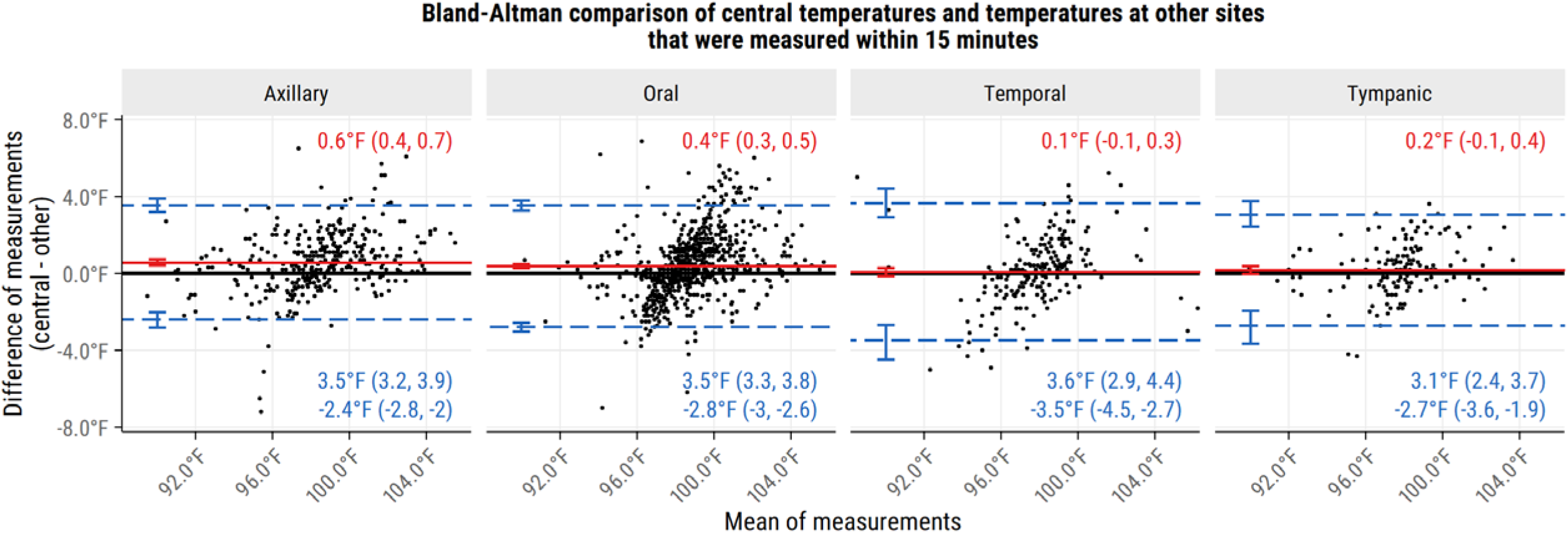
Bland-Altman comparison of central temperatures with temperatures taken at other sites within 15 minutes. Data are from adult critical care patients. All thermometer sites show similar, poor levels of agreement with central temperatures, though mean differences from central temperatures appear larger for axillary and oral thermometers than for temporal and tympanic thermometers. Confidence intervals are 95%. Mean differences (mean biases) are shown in red and limits of agreement (mean difference ± 2 standard deviations) are shown in blue. Central temperatures include pulmonary artery, esophageal, urinary bladder, rectal, and core.

**Figure 2.**
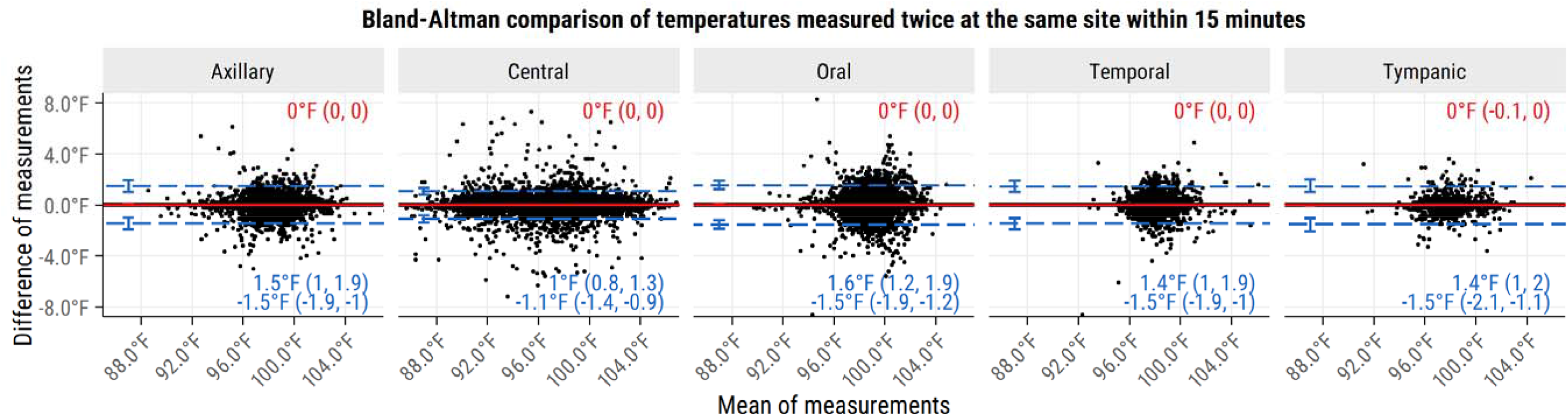
Bland-Altman comparison of temperatures taken twice at the same site with 15 minutes. The results show poor repeatability for all temperature sites, suggesting that a major reason for remeasurement may have been suspected clinician or patient error during a measurement. Additionally, measurements taken at central sites often reached low values that are generally inconsistent with life, but common for measurement technique errors. As previously, data are from adult critical care patients, mean differences are shown in red, limits of agreement are shown in blue, and confidence intervals are 95%.

## Discussion

Quickly retaken temperatures have very low repeatability and agreement in EHR, to a degree unprecedented by previous, non-EHR research on thermometer performance.^5,6^ Haimovich et al.^1^ observed this for TATs and we observed it for all major non-invasive thermometer types. Anomalously cold central temperatures were also common in both studies.^1^

A natural explanation is that clinicians often retook temperatures within minutes because of user or patient errors during measurement. Common errors include patient movement, inadvertent thermometer activation, and insufficiently deep rectal or esophageal probes. This last error produces artificially cold temperatures, but correcting it discomforts patients, so clinicians sometimes try non-invasive thermometers instead.

The explanation means that quickly retaken EHR measurements do not reflect device accuracy or precision in correct use. To detect similar problems in other EHR-based studies, researchers should place themselves in the clinicians’ shoes and carefully consider why actions listed in EHR were taken.

## Data Availability

All data used in this study are available online from PhysioNet as part of the eICU Collaborative Research Database, version 2.0. Given our conflicts of interest on this research topic, we have also provided code that can be used to check and replicate all study findings in the interest of transparency.

https://physionet.org/content/eicu-crd/2.0/

https://github.com/charlesharding/temp-retake-issues

## Acknowledgments

None.

## Online-only supplement

None. (Not allowed for the journal’s brief reports.)

## Code and data availability

For full transparency, all code and data for this study have been made available online. Please see the methods section for more details.

## References

1. Haimovich A, Taylor R, Krumholz H, Venkatesh A. Performance of temporal artery temperature measurement in ruling out fever: implications for COVID-19 screening. J Gen Intern Med. Published online 2020. doi:10.1007/s11606-020-06205-2

2. Pollard T, Johnson A, Raffa J, Celi LA, Badawi O, Mark R. ICU Collaborative Research Database (version 2.0). PhysioNet. Published online 2019. doi:https://doi.org/10.13026/C2WM1R

3. Scripts for analyzing quickly retaken temperatures in eICU datasets. Published 2020. Accessed November 14, 2020. https://github.com/charlesharding/temp-retake-issues

4. Pollard TJ, Johnson AEW, Raffa JD, Celi LA, Mark RG, Badawi O. The eICU collaborative research database, a freely available multi-center database for critical care research. Sci Data. 2018;5:18017. doi:10.1038/sdata.2018.178

5. Davie A, Amoore J. Best practice in the measurement of body temperature. Nurs Stand. 2013;24(42):42–49. doi:10.7748/ns2010.06.24.42.42.c7850

6. Jefferies S, Weatherall M, Young P, Beasley R. A systematic review of the accuracy of peripheral thermometry in estimating core temperatures among febrile critically ill patients. Crit Care Resusc. 2011;13(3):194–199.

